# The Histological Diagnosis of Colonic Adenocarcinoma by Applying Partial Self Supervised Learning

**DOI:** 10.1101/2020.08.15.20175760

**Authors:** Syed Usama Khalid Bukhari, Asmara Syed, Syed Khuzaima Arsalan Bokhari, Syed Shahzad Hussain, Syed Umar Armaghan, Syed Sajid Hussain Shah

**Affiliations:** Department of Computer Science, The University of Lahore, Islamabad, Pakistan; Department of Pathology, Faculty of Medicine, Northern Border University, Arar, Kingdom of Saudi Arabia; Doctors Hospital, Lahore, Pakistan; Electrical Engineering Department, National University of Technology, Islamabad, Pakistan; Biomedical Engineering, Riphah International University, Islamabad, Pakistan; Department of Pathology, Northern Border University, Arar, Kingdom of Saudi Arabia

**Keywords:** Colon, Adenocarcinoma, Artificial Intelligence

## Abstract

**Background:** The cancer of colon is one of the important cause of morbidity and mortality in adults. For the management of colonic carcinoma, the definitive diagnosis depends on the histological examination of biopsy specimens. With the development of whole slide imaging, the convolutional neural networks are being applied to diagnose colonic carcinoma by digital image analysis.

**Aim:** The main aim of the current study is to assess the application of deep learning for the histopathological diagnosis of colonic adenocarcinoma by analysing the digitized pathology images.

**Materials & Methods:** The images of colonic adenocarcinoma and non neoplastic colonic tissue have been acquired from the two datasets. The first dataset contains ten thousand images which were used to train and validate the convolutional neural network (CNN) architecture. From the second dataset (Colorectal Adenocarcinoma Gland (CRAG) Dataset) 40% of the images were used as a train set while 60% of the images were used as test dataset. Two histopathologists also evaluated these images. In this study, three variants of CNN (ResNet-18, ResNet-34 and ResNet-50) have been employed to evaluate the images.

**Results:** In the present study, three CNN architectures(ResNet-18, ResNet-30, and ResNet-50) were applied for the classification of digitized images of colonic tissue. The accuracy (93.91%) of ResNet-50 was the highest which is followed by ResNet-30 and ResNet-18 with the accuracy of 93.04% each.

**Conclusion:** Based on the findings of the present study and analysis of previously reported series, the development of computer aided technology to evaluate the surgical specimens for the diagnosis of malignant tumors could provide a significant assistance to pathologists.

## 1 Introduction

Cancers are an important cause of mortality and mortality in the developed countries as well as in the developing nations after the better control of infectious diseases. The estimated number of new cases of cancer is more than eighteen million with more than 9 million deaths in 2018 (1). The colorectal (colon and rectum) cancer ranked number 3 according to the number of malignant tumors and it is the 2^nd^ common cause of death due to cancer in both genders (male and female) all over the world (1). A difference in the occurrence of malignant tumors among the different countries has been documented in the literature (2). This variation in the prevalence rate of different types of cancers may be attributed to genetic and environmental factors.

A rising trend in the incidence of malignant tumors has been observes around the globe which may be attributed to the increase in the population as well as increase in the elderly population. Malignancy may occur at any age but most common histological types of cancer are diagnosed in old age. The most common age group is 51 – 60 years (3). It has been predicted that the death number may rise by sixty percent until 2035 (4).

The common clinical features of colorectal cancers include rectal bleeding, altered bowel habits, anemia and weight loss (5). Carcinoembryonic antigen is commonly used as a tumor marker and as a screening test but definitive diagnosis requires tissue biopsy for histopathological examination. The adenocarcinoma is the most common histological type of malignant tumor of colon and rectum. The adenocarcinoma is characterized by large neoplastic cells with nuclear pleomorphism and hyperchromasia. The tumor cells attempt to make glandular structures.

The important risk factors of colonic tumors are high intake of meat, old age, family history, obesity and smoking (6–7).

With the increase in exposure to number of risk factors, the incidence of colonic cancer is expected to rise which will generate more work load on the histopathologists. The application of smart technology such as artificial intelligence may be helpful in this regards. The aim of the present study is to assess the application of deep learning for the histopathological diagnosis of colonic adenocarcinoma by analysing the digital pathology images.

## 2 Materials & Methods

Total of ten thousand digital images, of histopathology slides, were obtained from LC25000 datasets. The data set is composed of 500 total images of colon tissue (250 benign colonic tissue and 250 colon adenocarcinomas) and augmented to 5,000 images of each using the Augmentor package [8]. As the dataset is augmented, mixed, and the author did not provide any information regarding augmentation; it was nearly impossible to split it into the train, and test sets as such. To use this dataset, we need to evaluate a different dataset for testing purpose. For this purpose, we used Colorectal Adenocarcinoma Gland (CRAG) Dataset [9]. The CRAG data set was developed for segmentation purpose, and in this study, we were working on a classification problem. Two histopathologists labeled and divided the CRAG dataset images into two categories, benign colonic tissue and colon adenocarcinomas. There were Forty-five (45) images of benign colonic tissues, and one hundred forty-eight (148) images of colonic adenocarcinoma. Three variants of ResNet architectures, ResNet-18, ResNet-34 and ResNet-50 were used to perform the classification (10). Following steps were taken to get the results.

## Step I

Using FastAI API (11), pretrain ResNet models (ResNet-18, ResNet-34, and ResNet-50) on ImageNet (12) were downloaded.

## Step II

LC25000 dataset was split into two sets, train with 80% of dataset and valid with 20% of the dataset. Train set contained 8000 images of both classes where valid set contained 2000 images of both classes.

All three ResNet variances were trained on LC25000 dataset using the learning rate: 1e-2, momentum = 0.9-0.8. To use both weights learned from ImageNet and our dataset; only weights of last ten layers were trained on LC25000 dataset.

## Step III

New labeled CARG dataset was split into two sets, train with 40%, and test with 60% of the dataset. Train set contained 78 images of both classes, where test set contained 115 images. This unusual split where test set is larger than the train set was done to show the performance of the self-supervised learning step, in which we trained our networks on LC25000. Reason to retrain the network is that our CRAG dataset resolution is different from the LC25000 dataset which is shown in figure 2.

**Figure 1.**
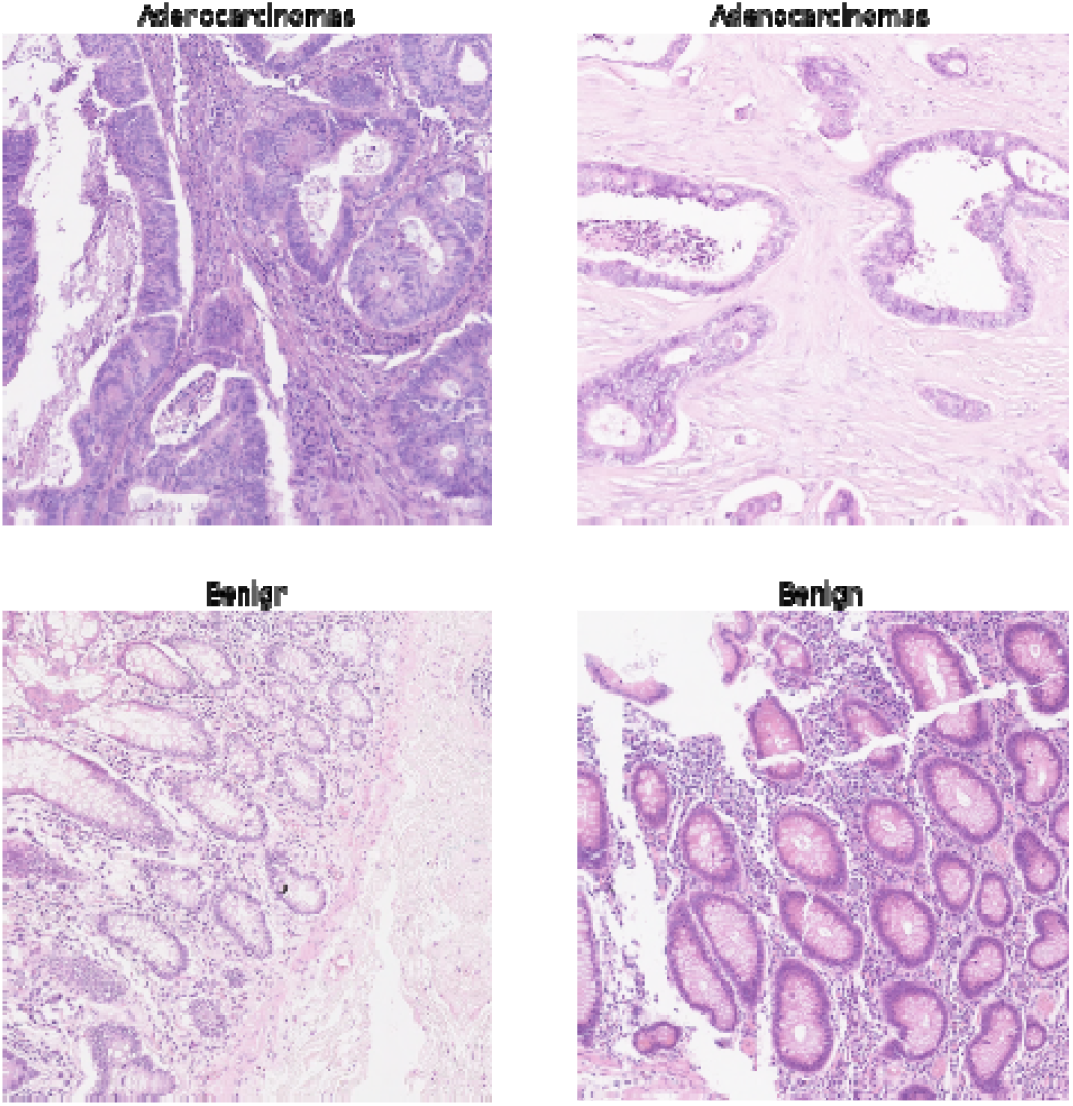
labelled CARAG dataset

**Figure 2.**
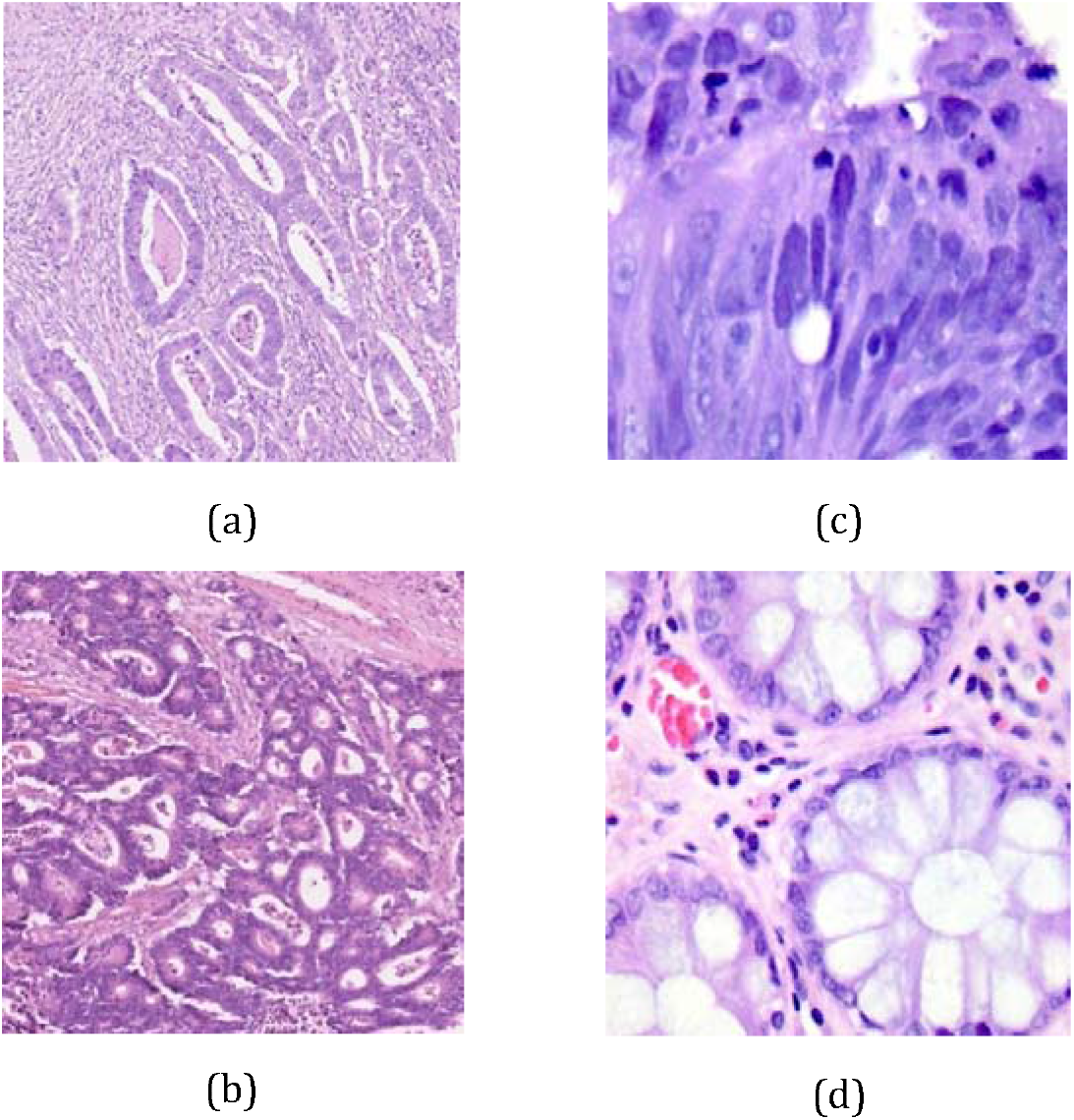
a and b belong CARG dataset, c and d belong to LC25000 dataset

All three ResNet variances were trained on new labeled CRAG dataset using the learning rate:1e-4, momentum = 0.9-0.8. To use both weights learned from self-supervised learning step and training set new labeled CRAG dataset; only the last two layers’ weights were adjusted.The results show that in scenarios, where we have a small dataset, and standard deep learning algorithm will end up over-fitting the data, the self-supervised learning step can be used.

## 3 Results

In the present study, three variants of convolutional neural networks (ResNet-18, ResNet-30, and ResNet-50) have been applied to assess the digital images for the diagnosis of colonic adenocarcinoma. The results of ResNet-50 revealed that there was discrepancy in the diagnosis in seven digital images with the diagnostic accuracy of 93.91% while in case of ResNet-18 and ResNet-30, the diagnostic discrepancy was in eight digital pathology images with a diagnostic accuracy of 93.04 % and 93.04% respectively. The results are shown in table 1 & 2.

**Table 1.**
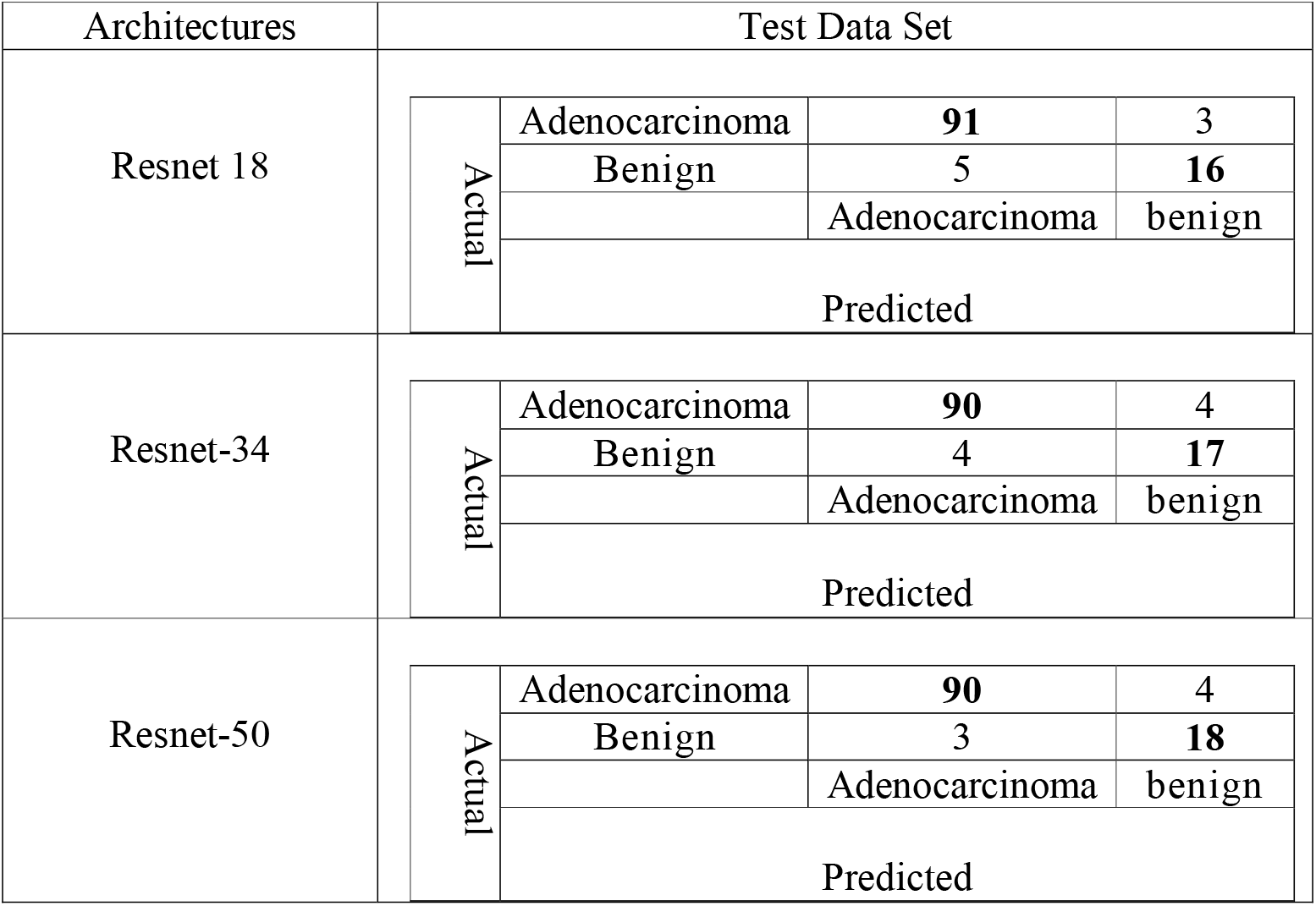
Confusion Matrix of Test Datasets.

**Table 2.** Detail Results of Test Dataset.

| MEASURE | RESNET-18 | RESNET-34 | RESNET-50 |
| --- | --- | --- | --- |
| SENSITIVITY | 94.79 | 95.74 | 96.77 |
| SPECIFICITY | 84.21 | 80.95 | 81.82 |
| PRECISION | 96.81 | 95.74 | 95.74 |
| NEGATIVE PREDICTIVE VALUE | 76.19 | 89.5 | 85.71 |
| FALSE POSITIVE RATE | 15.79 | 19.05 | 18.18 |
| FALSE DISCOVERY RATE | 03.19 | 04.26 | 04.26 |
| FALSE NEGATIVE RATE | 05.21 | 04.26 | 03.23 |
| ACCURACY | 93.04 | 93.04 | 93.91 |
| F1 SCORE | 95.79 | 95.74 | 96.26 |
| MATTHEWS CORRELATION COEFFICIENT | 75.94 | 76.70 | 80.01 |

## 4 Discussion

The analysis of results of the present study revealed that the diagnostic discrepancy of convolutional neural network (CNN) was less than 7% and the computer aided diagnostic system was able to diagnose and differentiate colonic adenocarcinoma from the non neoplastic colonic tissue in 93% of digitized pathology images of colonic tissue.

In a study conducted by Chen PJ et, the computer aided diagnostic system revealed 96.3% sensitivity with a negative predictive value of 91.5% for the detection of neoplastic or non neoplastic colorectal polyps (13). Another study performed on the image analysis of colorectal lesions by computer aided diagnostic system showed the sensitivity of 83.9 % and specificity of 82.6% (14). The accuracy of different deep learning models for the classification of colonic lesions varies from 80% to 99.2% (15–18). A study carried out by Chen M et al for the classification of benign and malignant tumors of liver by analysing the H&E images with the application of neural network revealed the accuracy of 96% (19).

In the present study, three CNN architectures(ResNet-18, ResNet-30, and ResNet-50) were applied for the classification of digitized images of colonic tissue. The sensitivity (96.77%) of ResNet-50 was the highest which is followed by ResNet-30 and ResNet-18 with the sensitivity of 95.74% and 94.79% respectively.

Recent advances in the artificial intelligence (AI) have revealed the astonishing performance of AI in many domains where it has surpassed human being (20). Since accurate diagnosis of the malignancy is of vital importance for the effective management of it. In spite of high clinical suspicion of malignancy, the final diagnosis can only be established after histopathological examination of the tissue. The surgical pathology is providing diagnostic services to the clinicians for the last hundred years. The key feature in the pathological evaluation for the diagnosis of cancers is the microscopic examination of tissue sections. The slides are made from tissue blocks and these slides are most commonly stained with hematoxylin and eosin stain. The basophilic structures of cells are stained with hematoxylin which appear as blue while eosinophilic structures take up the pinkish color after staining with eosin. The basophilic structures in the cells includes nucleus, rough endoplasmic reticulum and ribosomes while majority of extracellular structures are stained with eosin (21). Hematoxylin and eosin stain has been used for the last one century for the diagnosis of cancer (22). This stain is used to visualize the detailed feature of nuclei and cytoplasm which have got vital importance in the differentiation of different type of cells and classification of neoplastic lesions (22). Hyperchromasia and pleomorphism along with arrangement of malignant cells are characteristic features of colonic adenocarcinoma which provide diagnostic clue to the histopathologists. The development of deep learning (a subset of artificial intelligence) made a breakthrough in the analysis of digital images with extraction of characteristic features. Marked improvement has been made in the image analysis with the development of s convolutional neural network (23).

Based on the present study findings and previously reported observations, The development of computer aided technology to evaluate the surgical specimens for the diagnosis of malignant tumors is expected to provide a significant assistance to the pathologists.

One of the limitation of the present study is that the proposed system is not marking the segmented part of lesion which could further assist the pathologist. Further studies in this regards are recommended.

## Data Availability

Data is taken from the following references
Borkowski, Andrew A., et al. "Lung and Colon Cancer Histopathological Image Dataset (LC25000).” arXiv preprint arXiv:1912.12142 (2019).
Graham, Simon, et al. "MILD-net: minimal information loss dilated network for gland instance segmentation in colon histology images.” Medical image analysis . 2019;52 : 199-211.

## Funding

None

## Conflict of interest

None

## Notes

### Competing Interest Statement

The authors have declared no competing interest.

### Author Declarations

Research Ethical Committee of The University of Lahore gave approval.

